# The effectiveness of Cenobamate in patients previously treated with Vagus Nerve Stimulation for drug resistant epilepsy

**DOI:** 10.1101/2024.12.09.24318297

**Authors:** Lara Hogeveen, Benjamin Legros, Alfred Meurs, Veerle De Herdt, Mathieu Sprengers, Stephanie Hödl, Ann Mertens, Stefanie Gadeyne, Robrecht Raedt, Frank Dewaele, Jelle Vandersteene, Paul Boon, Kristl Vonck

## Abstract

**Background:** Vagus Nerve Stimulation (VNS) is an efficacious neuromodulatory treatment for patients with drug resistant epilepsy (DRE). In many VNS treated patients, seizures are not fully controlled. These patients often represent a highly refractory group in whom novel anti-seizure medication (ASM) may be tried. This study evaluates the effectiveness of Cenobamate (CNB) in patients previously treated with VNS.

**Methods:** The medication history of all patients treated with VNS at Ghent University Hospital and HUB-Hôpital Erasme Brussels between 1995 and 2024 was reviewed. In patients who received an add-on treatment with CNB after at least 12 months of VNS therapy, mean monthly seizure frequency before VNS, after VNS, and after CNB was assessed.

**Results:** 54/620 patients treated with VNS between March 1995 and November 2023 were treated with CNB after a median VNS treatment time of 9 years. With VNS, 13/52 (25%) had a seizure frequency reduction of ≥50%. Side-effects were mild and stimulation-related. After add-on of CNB (median daily dosage of 200mg) for a median follow-up of 10 months, 24/54 (44.4%) became responders. Of these patients, 6 (11.1% from total cohort) reported seizure freedom for a median duration of 26 weeks. Almost half (46.2%) of the VNS non-responders became CNB responder. The median number of concomitant ASM was 3. In 20/54 (37%), the number of ASM was reduced. 7/54 (13%) stopped CNB because of side-effects. Fatigue was the most prevalent side effect in 23/54 patients (42.6%).

**Conclusion:** In DRE patients treated with VNS therapy who could benefit from further improvement in seizure control, add-on treatment with CNB is useful. This most recently marketed ASM leads to a significant improvement in seizure frequency in more than 40% of the patients.

## Introduction

About 30% of patients with epilepsy (PwE) do not achieve seizure freedom after trials with two or more well-chosen and well-tolerated ASM and are considered to have drug resistant epilepsy (DRE) (1). Patients with DRE should be referred to a specialized epilepsy center for advanced diagnostics and treatment, where suitability for epilepsy surgery or neuromodulation therapies such as Vagus Nerve Stimulation (VNS) and Deep Brain stimulation (DBS) can be evaluated.

VNS was approved 30 years ago and is the most widely used neuromodulation therapy for DRE (2–4). Action potentials are induced in the afferent vagal myelinated A- and B-fibers that project to the brainstem nucleus of the solitary tract, then to the locus coeruleus and parabrachial nucleus leading to the release of neurotransmitters in brain structures such as the limbic system, thalamus, and cerebral cortex. Different neurotransmitters are thought to be involved in the mechanism of action, including norepinephrine, serotonin, Ach, GABA and aspartate (5,6). VNS-induced catecholamine release from the locus coeruleus in the brainstem plays a pivotal role in VNS control over epileptic activity (5,6).

Several studies have demonstrated the efficacy of VNS over the first year in improving both seizure frequency and severity when concomitant ASM remain unchanged, with an increasing efficacy of VNS with prolonged duration of the therapy. Responder rates, defined as the percentage of patients receiving a reduction in seizure frequency of 50% or more, have been reported as 56.9%, 62.8% and 64.1% at 12, 18 and 24 months respectively (7–9). Morris et al. found that after one year open-label follow-up, 37% of patients showed a seizure reduction of ≥50% (10). After long-term follow-up, this number increased up to 40-65% (10–15). Long-term data of effectiveness most likely represent a combined therapy approach including ASM, VNS and other previously installed anti-epileptic therapies. PwE who receive VNS treatment often represent a highly refractory group (8,10,16–19). With VNS, a limited number of patients become seizure free and even more than one-third (36.3%) of patients experience less than 50% reduction in seizure frequency, classifying them as non-responders (18). For these patients, trials with newly developed treatment options remain an option.

Cenobamate (CNB) is a relatively new ASM that received FDA approval in 2019 and EMA approval in 2021 for focal seizures. The mechanism of action is multifaceted, targeting multiple pathways. It involves the inhibition of voltage-gated sodium channels, including both transient and persistent sodium currents. Additionally, it modulates GABA-A receptors through allosteric binding, enhancing the receptor’s response to GABA (20,21). Several randomized controlled trials (RCTs) have demonstrated a responder rate of around 50% and seizure freedom rates of more than 20% (22–24). Treatment with CNB requires dedicated management by neurologists due to its pharmacokinetic and pharmacodynamic profile but is often well tolerated with side effects diminishing over time and long-term retention rates are high (22,25–27). In this study, we investigated the effectiveness of CNB as an adjunctive ASM in a large group of DRE patients who had previously been treated with VNS and other ASM.

## Materials and methods

### Data extraction

In this retrospective multicenter study, the medication history of all PwE treated with VNS between March 1995 and May 2024, at Ghent University Hospital and HUB-Hôpital Erasme Brussels were reviewed. Adults who received treatment with CNB after at least 12 months of VNS treatment and had at least one follow-up visit after the start of CNB were included. Patients on a daily CNB dose of 12.5mg at the last follow-up visit were excluded from further analysis.

Ghent University hospital acted as data collector. Clinical data including age, sex, epilepsy type, duration of epilepsy, lifetime and concomitant ASM and other anti-epileptic therapies were assessed. Dose and treatment duration of CNB from subjects who continued treatment, VNS treatment duration and VNS parameters at the last follow-up visit were determined. Monthly seizure frequency before and after VNS and before and after add-on of CNB were collected; seizure frequency was retrospectively collected from patient files based on patient self-reports. The data is subjective in nature, as no seizure diaries or systematic recording methods were utilized.

The mean monthly seizure frequency before VNS refers to the mean monthly seizure frequency over the last three months before the start of VNS treatment. Post-VNS therapy, we investigated the mean monthly seizure frequency after one year of VNS during the last three months of the first treatment year. This was considered a significant time point for evaluating the efficacy of VNS, given that its effectiveness tends to improve over time (19,28,29). Subsequently, we assessed the mean monthly seizure frequency at maximum follow-up period after VNS, which refers to the mean monthly seizure frequency during the last three months of VNS before CNB association. Mean monthly seizure frequency at the maximum follow-up with CNB was recorded at the last follow-up visit and was dependent on the duration of the follow-up. Seizure freedom with VNS was determined during the one-year follow-up consultation and at the maximum follow-up prior to CNB association. Seizure freedom with CNB was evaluated at the last follow-up visit.

### Outcome parameters

The effectiveness of VNS and CNB was evaluated through two primary metrics. The median reduction in mean monthly seizure frequency, calculated and reported as an absolute value and the 50% responder rate which represents the percentage of patients achieving a seizure frequency reduction of at least 50%. Reported seizure freedom, including its duration, was recorded. We assessed seizure freedom with CNB according to the ILAE definition, which considers a patient seizure free if they experience no seizures for at least three times the length of their longest pre-treatment inter-seizure interval In the preceding 12 months (1). For VNS, the median reduction in mean monthly seizure frequency was determined by comparing seizure frequency before VNS to after one year of treatment, as well as to the maximum follow-up period.

### Seizure frequency reduction was calculated using the following formulas

- (mean monthly seizure frequency after 1 year of VNS – mean monthly seizure frequency before VNS) / mean monthly seizure frequency before VNS x100.
- (mean monthly seizure frequency after maximum follow-up of VNS – mean monthly seizure frequency before VNS) / mean monthly seizure frequency before VNS x100.

A responder at one year was defined as a patient with a seizure frequency reduction of 50% or more after one year of VNS therapy compared to before VNS. A responder at maximum follow-up achieved a seizure frequency reduction of 50% or more compared to before VNS.

For CNB, the median reduction in mean monthly seizure frequency was determined by comparing seizure frequency at maximum follow-up with CNB to the seizure frequency at the maximum follow-up with VNS corresponding to the period immediately before CNB was added.

### Seizure frequency reduction was calculated using the following formula

- (mean monthly seizure frequency after maximum follow-up of CNB – mean monthly seizure frequency after maximum follow-up of VNS) / mean monthly seizure frequency after maximum follow-up of VNS x100.

A responder to CNB therapy was defined as a patient achieving a seizure frequency reduction of 50% or more at the last follow-up visit compared to the maximum follow-up with VNS. Additionally, to examine tolerability, adverse events were recorded descriptively during consultations based on patient self-reports. The retention rate for CNB was assessed as the percentage of patients continuing CNB treatment at the last visit. Post-hoc sub-analyses were conducted to examine specific subgroups with the aim of identifying characteristics that could explain the observed effects. Subgroups included: VNS non-responders, CNB responders, seizure-free patients and patients with intolerance to CNB.

### Statistics

Data were entered into an Excel database and analyzed using IBM SPSS Statistics version 28.0.1.1.1. Due to the small sample size, only descriptive statistical analyses were performed. Median values, along with the interquartile range and range, were reported due to the high variability observed in most parameters. Correlations, along with confidence intervals, were calculated using the Spearman’s coefficient to identify a potential relationship between variables and to understand the strength and direction of these relationships. A complete-case analysis was performed. Missing values were due to the unavailability of older data in patient files or the absence of patient-reported documentation of specific information, reflecting the retrospective study design.

## Results

### Demographics and clinical characteristics

620 PwE were treated with VNS in two Belgian epilepsy centers between March 1995 and May 2024. 54/620 (30/54 Ghent University Hospital, 24/54 Erasmus University Hospital Brussels) patients (age 18-74 y, median: 38, M/F 26/28) were additionally treated with CNB after at least 12 months of VNS treatment. In 8 patients VNS had been turned off or removed before CNB association due to ineffectiveness or side effects. One patient died of an underlying disease 5 months after CNB association. Characteristics of the patients can be found in table 1. 83.3% of patients had focal epilepsy. The median duration of epilepsy was 28 years (IQR 20.2, range 10-71), the median number of ASM patients received before CNB association was 10 (IQR 4, range 5-18), the median number of seizures in the last three months before VNS was 17.8 (IQR 25.4, range 2-173) (N=48). Between one year of VNS treatment and the association of CNB, therapy changes were made in 43 patients.

**Table 1:** Clinical and treatment characteristics. Abbreviations: CNB, Cenobamate; ASM, anti-seizure medication; VNS, Vagus Nerve Stimulation; IQR, interquartile range; DBS, Deep Brain Stimulation.

| Clinical and treatment characteristics | Details |
| --- | --- |
| Sex, n male (%) | 26/54 (48.1) |
| Median age at start CNB, years (IQR, range) | 38 (20.7, 18-74) |
| Epilepsy type, n (%) |  |
| - Focal | 45/54 (83.3) |
| - Multifocal | 5/54 (9.3) |
| - Generalized | 3/54 (5.6) |
| - Combined | 1/54 (1.9) |
| Median duration of epilepsy at start CNB, years (IQR, range) | 28 (20.2, 10-71) |
| Median number of life-time ASM (IQR, range) | 10 (4, 5-18) |
| Median number of monthly seizures before VNS implantation (IQR, range) | 17.8 (25.4, 2-173) |
| Non-pharmacological treatments prior to VNS, n (%) |  |
| - Surgery | 7/54 (13) |
| - Radiotherapy (surgery or ablation) | 4/54 (7.4) |
| Therapy changes between one year post-VNS and before CNB start, n (%) |  |
| - ASM trial | 40/54 (74.1) |
| - Ketogenic diet trial | 4/54 (7.4) |
| - DBS implantation | 5/54 (9.3) |
| - Surgery | 2/54 (3.7) |
| - No changes | 11/54 (20.4) |
| Median number of concomitant ASM at start of CNB addition (IQR, range) | 3 (1, 2-6) |
| Median number of concomitant ASM at last CNB follow-up visit (IQR, range) | 3 (1.3, 1-5) |
| Median dose of CNB in mg at last CNB follow-up visit (IQR, range) | 200 (100, 50-350) |
| Median VNS treatment duration in years before CNB association (IQR, range) | 9 (11, 1-26) |
| Median CNB duration in months up to the last visit (IQR, range) | 10 (5, 1-17) |

Median VNS treatment duration before CNB association was 9 years (IQR 11, range 1-26). Median VNS parameters, of patients whose VNS was turned on, at last visit were as followed: output of 2 mA (IQR 1, range 0.38-2.75), frequency of 20Hz (IQR 10, range 10-30), pulse width of 250 µsec (IQR 250, range 130-500), duty cycle of 10% (IQR 15, range 5-45) (N=46). At start of CNB treatment the median number of concomitant ASM was 3 (IQR 1, range 2-6). The median follow-up period of CNB association was 10 months (IQR 5, range 1-17). The median dose of CNB at last visit, from the subjects who continued CNB, was 200 mg (IQR 100, range 50-350). 33/54 (61.1%) patients used a daily dose of 200mg or more.

### Vagus Nerve Stimulation effectiveness

Data on seizure frequency was collected for two follow-up periods. At the 1 year follow-up, information was available in 46/54 patients. The responder rate was 30.4% (14/46). Of these, 1 patient (2.2% from total cohort) reported seizure freedom, for a duration of 4 months but her VNS was in the off-mode. The median reduction in seizure frequency was 0% (IQR 50, range 0-100).

Meanwhile, after a median VNS treatment duration of 9 years (IQR 11, range 1-26) prior to CNB association, data regarding seizure frequency was available in 52/54 patients. The responder rate was 25% (13/52). None of the patients reported seizure freedom at maximum follow-up with VNS treatment. The median seizure frequency reduction was 0% (IQR 49, range 0-93) (N=48).

### Cenobamate effectiveness

After a median CNB treatment duration of 10 months (IQR 5, range 1-17), the median reduction in seizure frequency was 32% (IQR 86.3, range 0-100). The responder rate was 44.4% (24/54). Of these, 6 patients (11.1% from total cohort) achieved seizure freedom at the last follow-up visit with a median seizure freedom duration of 26 weeks (IQR 35, range 4-60). The CNB retention rate was 87%. None of the patients discontinued treatment because of ineffectiveness alone.

### Concomitant anti-seizure medications

All patients used concomitant ASM. At the start of the treatment with CNB the median number of concomitant ASM was 3 (IQR 1, range 2-6). This number remained unchanged at maximum follow-up with CNB (IQR 1.3, range 1-5). However, in 20/54 (37%) patients, the number was decreased with 1 ASM at maximum follow-up. The most frequently used ASM at last follow-up visit i.e. in >10% of patients, are presented in table 2. Brivaracetam was most frequently used as concomitant ASM, followed by Lacosamide, Topiramate and Carbamazepine.

**Table 2:** number of patients (in %) using ASM at last follow-up visit of CNB (ASM used in more than 10% of patients).

| ASM | % in total population | % in CNB responders | % in seizure free group | % in subgroup with intolerance |
| --- | --- | --- | --- | --- |
| Brivaracetam | 44.4 | 41.7 | 83.3 | 47.2 |
| Lacosamide | 33.3 | 29.7 | 16.7 | 36.1 |
| Topiramate | 31.5 | 41.7 | 66.7 | 36.1 |
| Carbamazepine | 31.5 | 20.8 | 33.3 | 20.4 |
| Valproic acid | 25.9 | 41.7 | 50.0 | 30.6 |
| Clobazam | 22.2 | 33.3 | 50.0 | 22.2 |
| Lamotrigine | 22.2 | 12.5 | / | 16.7 |
| Levetiracetam | 22.2 | 20.8 | 16.7 | 27.8 |
| Clonazepam | 16.7 | 12.5 | / | 13.9 |
| Pregabalin | 14.8 | / | / | 13.9 |
| Perampanel | 11.1 | / | / | / |
| Gabapentin | / | / | 16.7 | / |

### Cenobamate tolerability

36/54 patients (66.7%) reported at least one side effect and CNB was discontinued in 7/54 patients (13%) because of this. The most frequently reported side effects can be found in table 3. All patients who stopped CNB reported having side effects, most frequently fatigue.

**Table 3:**
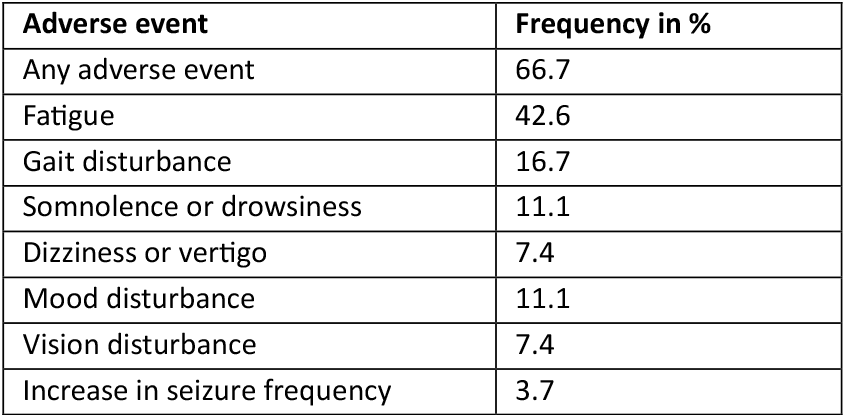
frequency of adverse events reported after add-on of CNB.

### Subgroup analyses

#### VNS non-responders

At maximum VNS follow-up, 39/52 (75%) patients were non-responders to VNS, with a median follow-up period of 12 years (IQR 10.8, range 1-26). There was no gender predominance, the median age was 35 years (IQR 19, range 18-74), median duration of epilepsy 28 years (IQR 19, range 10-71) and the median number of life time ASM 10 (IQR 3.5, range 5-18). The responder rate after CNB addition in this group was 46.2% (18/39) after a median follow-up of 10 months (IQR 7, range 3-17). Of these 18 responders, 5 patients (12.8% from total cohort) reported seizure freedom at last follow-up visit for a median time of 20 weeks (IQR 42, range 4-60). For the 6^th^ seizure free patient, VNS effectiveness data was not available at last follow-up visit.

#### CNB responders

In the 24/54 patients who were responder to add-on therapy with CNB, for a median duration of 9.5 months (IQR 8.9, range 3-17), the median age was 33 years (IQR 21, range 18-62). 14/24 patients (58.3%) were male with focal onset epilepsy in 21/24 patients (87.5%), the other patients had multifocal epilepsy. The median number of life time ASM was 9.5 (IQR 4, range 5-18). The median time of VNS treatment before CNB association was 6.3 years (IQR 8, range 2-26).

The median dose of CNB in this group was 200mg (IQR 118.8, range 50-350). 17/24 (70.8%) patients were on a daily dose of 200mg or more and 5/24 (20.8%) used a daily dose of at least 300mg. Median VNS parameters included an output current of 2 mA (IQR1, range 0.75-2.75), frequency of 30Hz (IQR 10, range 10-30), pulse width of 500 µsec (IQR 250, range 130-500), duty cycle of 10% (IQR 15, range 10-39). In 3 patients VNS had been turned off before CNB association.

Data on seizure frequency reduction after one year of VNS was available in 19/24. The responder rate was 42.1% (8/19). Data on seizure frequency reduction after maximum follow-up of VNS was available in 22/24. The responder rate was 18.2% (4/22) after a median VNS follow-up time of 6.3 years (IQR 8, range 24). Earlier non-pharmacological treatments before CNB in this group included ANT-DBS (N=2), surgery (N=3) and radiotherapy (surgery or ablation) (N=3). The median number of concomitant ASM at last visit was 3 (IQR 2), which did not differ from the total population. Brivaracetam, Topiramate and Valproic acid were the most frequently used ASM, followed by Clobazam (table 2).

#### Seizure free with CNB

Investigation of the characteristics of the 6 seizure free patients after CNB association for a median time of 14 months (IQR 7.4, range 4-17), showed no gender predominance, a median age of 46 years (IQR 37, range 25-62) and focal onset epilepsy in all patients. The median duration of epilepsy was 35.5 years (IQR 37, range 19-62). 2 patients had previous non-pharmacological treatments, which included surgery (N=1), radiosurgery (N=2) and DBS (N=1). The median daily dose of CNB was 150mg (IQR 175, range 50-300). 3/6 patients (50%) used a dose of 200mg or more. The median number of concomitant ASM did not differ from the total population; at start median 3 (IQR 2.5, range 2-6), at the last visit median 3 (IQR 3, range 2-5). In 2/6 (33.3%) stopping 1 ASM was possible. The most frequently concomitantly used ASM was Brivaracetam, followed by Topiramate (Table 2).

Data on seizure frequency reduction after one year of VNS was available in 5/6 patients. 2/5 (40%) were responders at this time. All seizure free patients with available data were non-responders to VNS treatment at the maximum follow-up period of 12 years (IQR 12, range 3-26).

Median VNS parameters included an output current of 2.25 mA (IQR 0.63, range 1.5-2.5), frequency of 25 Hz (IQR 10, range 20-30), pulse width of 500 µsec (IQR 250, range 250-500), duty cycle of 25% (IQR 25, range 10-35). In 1 patient VNS was turned off before CNB association.

#### Intolerance to CNB

The characteristics of the 36/54 patients who reported adverse events on CNB were analyzed. The median age in this group was 39.5 (IQR 23.8, range 54), with a female predominance (52.8%). The median dosage of CNB from patients who continued CNB was 250mg (IQR 75, range 300). 23/36 (63.9%) patients were on a dose of 200mg or more, 7/36 (19.4%) were on a dose of 300mg or more. 7/36 (19.4%) patients stopped CNB. The median duration of CNB treatment up to the last visit was 10 months (IQR 4.4, range 16). The median number of concomitant ASM did not differ from the total group. 3 (IQR 1, range 4) at the start and at the end (IQR 2, range 4).

Brivaracetam was most frequently used in this group, followed by Lacosamide and Topiramate (see table 2). Median VNS parameters were as follows: output of 2 mA (IQR, range), frequency of 20Hz (IQR, range), pulse width of 500 µsec (IQR, range), duty cycle of 10% (IQR, range).

## Discussion

This retrospective study aimed to evaluate the potential benefit of CNB as adjunctive therapy in patients with DRE previously treated with VNS. VNS is a key therapeutic neuromodulation option for DRE but a subset of patients continues to be non-responsive even after prolonged VNS treatment. In these patients, re-evaluation and consideration of recently developed therapies, such as CNB, is necessary. The key question is whether introducing a novel ASM can yield additional improvement in this refractory group already treated with neuromodulation. By analyzing clinical data from Ghent University Hospital and HUB-Hôpital Erasme Brussels, we assessed the effectiveness, tolerability, and characteristics of clinical outcome associated with the introduction of CNB.

The analysis of CNB add-on in 54 VNS-treated patients who could benefit from therapeutic improvement according to best medical practice revealed a significant seizure frequency reduction of 32%. Notably, 44% of the patients became responders following the addition of CNB. The patients in this study had a median duration of epilepsy of 28 years and a lifetime use of 10 ASM at the time of CNB initiation, demonstrating its effectiveness in a severely refractory population. Although this is a refractory group, almost half of the VNS non-responders (46.2%), became responders following the addition of CNB and 15.4% of the VNS non-responders reported seizure freedom. Several studies point towards both a high response rate and seizure freedom rate with CNB in highly refractory epilepsy patients (30,31).

In comparison to other studies investigating CNB as an adjunctive treatment, responder rates vary between 57% and 81%, with follow-up periods ranging from 9 weeks to 30 months (26,27,32). Abou-Khali et al. reported a 26.7% seizure freedom rate for at least 12 months in a post-hoc subgroup analysis of 45 patients previously treated with VNS/RNS or epilepsy surgery (31). Our relatively lower responder rates likely reflect the high refractoriness of our cohort. Reported seizure freedom rates at last follow-up visit in our study (13%) were consistent with earlier real-world studies where it ranges from 5.3% to 14%, though it is unclear whether the ILAE definition for seizure freedom was consistently used in these studies (23,24,27,33).

The patient group in our study showed a low response to VNS with a median responder rate of 25% after a median therapy duration of nine years. On the other hand, 42.1% of patients in the CNB responder group were also responders to VNS after one year of therapy. Predicting outcomes based on early VNS effectiveness remains challenging, as no significant correlations were identified. The overall VNS responder rates in the literature typically range from 45% to 65% (10–15). Again, the lower numbers in our population likely reflects the highly refractory nature of the patient cohort in this multicentric study. According to best medical practice, treating physicians likely prioritized introducing CNB to the most refractory patients, such as VNS non-responders, in an effort to improve seizure control when novel therapies became available. This was the case in 75% of the total study population versus 25% of patients who were VNS responders but in whom there was still room for improvement. These findings suggest that adding CNB after neuromodulation may still improve outcomes in some patients and should be considered (30). 36.1% of VNS non-responders did not respond to CNB, emphasizing the need for alternative treatment strategies in this subgroup.

In our responder group, Brivaracetam was the most frequently used ASM, followed by Topiramate. Interestingly nearly all seizure-free patients were on Brivaracetam, indicating a potentially favorable combination with CNB. Other studies have reported higher proportions of seizure-free patients with concomitant Levetiracetam or Zonisamide use (33). Brandt et al. reported a higher seizure freedom rate with CNB administration compared to placebo, regardless of the concomitant ASM used. However, at a daily dose of 400 mg CNB, the median seizure reduction was 89.0% in patients using non-sodium channel blockers, compared to 54.3% in those using sodium channel blockers (34). Combining VNS with specific ASM offers benefits beyond efficacy, including reduced side effects and improved tolerability. VNS is known for positive effects such as enhanced alertness, mood, and cognition, potentially supporting ASM adherence (35–39). In our study, 66.7% of patients reported at least one side effect after a median follow-up of 10 months—a rate slightly lower than the 68.2–74.1% reported in previous studies over 3 to 12 months (26,27,40). Sedation-related effects, such as fatigue, somnolence, and drowsiness, were most common, consistent with prior findings (31,41,42). Somnolence was reported in only 11.1% of patients, significantly lower than the 28.1–64.9% reported in earlier research (27,40,43–45).

Polytherapy remains challenging in epilepsy management, but dose adjustments when adding CNB can optimize outcomes (27,40,43–45). Brivaracetam and Topiramate were associated with improved outcomes but also higher rates of sedative side effects in our cohort. Among patients reporting sedation, 50% used Brivaracetam, often alongside Lacosamide, Topiramate, and Valproic Acid, each present in over 30% of cases. Lacosamide was linked to side effects without proportional efficacy benefits. Expert opinions recommend reducing Lacosamide during CNB titration to minimize pharmacodynamic interactions (46). Similarly, Clobazam increases CNB serum levels, potentially enhancing both efficacy and fatigue risk (47). Brandt et al. also noted increased dizziness when CNB was combined with sodium channel blockers (34).

Reducing ASM burden appears feasible with VNS and CNB. In our cohort, 37% of patients discontinued at least one ASM, consistent with studies reporting that 50% or more reduced drug burden after successful CNB use (22,48).These findings highlight the importance of optimizing polytherapy regimens to balance efficacy and tolerability.

The retention rate for CNB in our study was approximately 87% after a median of 10 months, comparable to other studies reporting retention rates between 80.4% and 89% with follow-up periods of 6.7 to 33 months (26,27,31,45,49). Variability in these outcomes may be attributable to differences in concomitant ASM use, CNB dosages, and follow-up periods. Notably, Abou-Khalil found that retention rates were higher in patients with VNS compared to those without, with rates of 80% in the VNS group (with or without surgery) versus 72.3% in patients without VNS. These findings suggest that VNS could positively influence CNB adherence and tolerability (31).

Side effects of CNB are well-documented and typically occur more frequently at higher doses (27,34). In our study, the median CNB dose among patients experiencing side effects but continuing treatment was 250 mg. Notably, 63.9% (23/36) of these patients were on a dose of 200 mg or more, and 19.4% (7/36) were on a dose of 300 mg or more. This distribution did not significantly differ from the total study population. Interestingly, no substantial differences in CNB dosage were observed between the total population and both responder and seizure-free groups. A median dose of 200 mg/day proved effective for seizure reduction, consistent with prior studies demonstrating CNB effectiveness even at lower doses without a clear dose-dependent effect (22,40,50). Nonetheless, over 70% of responders in our cohort were on doses of at least 200 mg, confirming this is a critical threshold. One in three patients were on a dose of 250 mg/day or more. This observation raises the hypothesis that refractory patients may benefit from higher CNB doses, potentially facilitated by improved tolerability due to combination therapies such as VNS (30). On the other hand, synergistic effects of combination therapies may allow some responders to achieve effectiveness at lower doses. Pena-Ceballos et al. found that among treatment responders, 67.4% was treated with CNB doses of ≥250 mg/day, which is higher than observed in our population (27).

In our total population, the median CNB dose at the last visit for patients continuing therapy was 200 mg/day, aligning with the target dose known to offer the optimal balance between efficacy and tolerability (22). In our cohort, 61.1% (33/54) of patients were on doses of 200 mg or more, and 19.4% (7/54) were on doses of 300 mg or more. These findings are comparable to, yet distinct from, previous studies. Steinhoff et al. reported that 58% of patients were on doses between 200 and 300 mg, while 11.2% exceeded 300 mg at their last visit (22), slightly lower than the proportions observed in our study. Another trial reported a mean CNB dose of 254 ± 82.1 mg/day, compared to our mean dose of 201 ± 74.9 mg/day.

While our findings provide valuable insights into the effectiveness and safety of CNB combined with VNS, several limitations must be acknowledged. The retrospective, real-world nature of the study introduces potential bias, which may limit generalizability. The small sample size, particular in subgroup analyses, and reliance on descriptive statistics prevent that definitive conclusions can be drawn. This study serves as a trend analysis, forming a foundation for future research. Our cohort’s lower VNS response rates may reflect inherent selection bias, as our centers predominantly treat patients with highly refractory epilepsy.

The lack of controlled conditions could introduce confounding factors, as therapy adjustments made alongside CNB initiation may have influenced outcomes. Moreover, seizure frequency and adverse event reporting are inherently subjective and the relatively short follow-up period for CNB—up to 17 months—may limit the assessment of long-term effectiveness and safety.

However, when focusing on patients with a minimum follow-up of six months, we observed a responder rate of 42.9%, suggesting a promising short-term benefit of CNB in this cohort. To strengthen evidence for the CNB and VNS combination and possible correlations, prospective trials with larger cohorts and controlled designs are needed. Such studies would validate our findings and provide robust conclusions on effectiveness and safety. Further investigation into the mechanisms underlying their additive effects is also warranted. CNB’s pharmacological actions—modulating voltage-gated sodium channels and enhancing GABA transmission—likely complement VNS’s neuromodulatory effects, but further research is required to elucidate these interactions.

## Conclusion

Our findings indicate that the addition of CNB to VNS therapy offers a promising treatment strategy for patients with DRE, particularly for those who remain refractory in spite of prolonged VNS therapy. In our cohort, 44.4% of patients achieved significant improvement in seizure control, with a retention rate of 87% after a median follow-up of 10 months. Even in this highly refractory group, 42.1% of the CNB responders, had also been responder to VNS at one-year follow-up. Remarkably, 46.2% of VNS non-responders—with a median VNS treatment duration of 12 years—became responders to CNB therapy, with 12.8% achieving seizure freedom.

Furthermore, the introduction of CNB was associated with a reduction in total ASM burden in over 30% of patients. These results emphasize the potential of CNB as an effective treatment even in highly refractory patients in whom there is room for improvement. It is crucial for clinicians to re-evaluate non-responders or partial responders to VNS when new ASM become available. Additionally, further research is needed to explore innovative neuromodulation techniques and refine strategies for this challenging population.

## Data Availability

All data produced in the present study are available upon reasonable request to the authors.

